# Identifying Explosive Epidemiological Cases with Unsupervised Machine Learning

**DOI:** 10.1101/2020.05.17.20104661

**Authors:** Serge Dolgikh

## Abstract

An analysis of a combined dataset of epidemiological statistics of national and subnational jurisdictions, aligned at approximately two months after the first local exposure to Covid-19 with unsupervised machine learning methods such as PCA and deep autoencoder dimensionality reduction allows to clearly separate milder background cases from those with more rapid and aggressive onset of the epidemics. The analysis and findings of this study can be used in evaluation of possible epidemiological scenarios and as an effective modeling tool to design corrective and preventative measures to avoid developments with potentially heavy impact

## 1 Introduction

An analysis of factors that can influence the course of the development of the epidemics in a given jurisdiction is both a challenging and interesting undertaking given the number of potential factors and their interaction. For example, a possible link between the effects of Covid-19 pandemics and a number of epidemiological factors including universal immunization program against tuberculosis with BCG vaccine was proposed in Miller et al. [1] and further investigated in [2-4]. Other factors, such as: gender and ethnicity; age demographics; social habits such as smoking; and others were investigated in a number of studies [5,6] and many others.

However, given the large quantity of factors that may have influence on the outcome of the epidemics in each case, identification of the most influential ones may represent certain challenge due to the number, complexity and interaction of contributing factors. In this work we attempt an analysis of the combined dataset of national and subnational reporting jurisdictions adjusted and aligned at the same time point of approximately two months after the first local exposure to the Covid-19 epidemics with the methods of unsupervised machine learning.

The unsupervised dimensionality / redundancy reduction methods such as Principal Component Analysis (PCA) [7] and unsupervised deep artificial neural network models such as autoencoders (AE) [8] allow to analyze the distribution of case data points in the informative parameter spaces identified by these methods and to attempt and in many instances, identify characteristic regions associated with the variable of interest, such as in this work, the severity of the epidemiological scenario in the jurisdiction. Establishing combinations of the latent and observable parameters that identify such regions can be used to evaluate and predict the risks of heavier epidemiological impacts in the jurisdiction proactively with the opportunity to make necessary corrections before the explosive onset of the epidemics would cause heavy costs to the society.

## 2 Method

The methodology is based on processing the input data expressed as a set of observable parameters that were identified and described in the study with unsupervised machine learning methods to identify and extract a smaller set of informative features. In many cases, evaluating distributions of data in the representations of informative components such as principal components in PCA or dimensionality reduction with neural network autoencoder models allows to identify and separate characteristic classes of cases in the observable data by essential latent parameters that can be linked to the observed outcome.

### 2.1 Combined Case Dataset

Timing considerations can be critical in the analysis of the development of an epidemiological scenario. For this reason, an effort was made to ensure that the data analyzed was recorded at a similar phase in the development of the epidemics in the jurisdiction. To clarify and emphasize the need for synchronization of epidemiological data, the zero time of the start of the global Covid-19 pandemics was defined in [2] as:

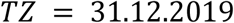

Evidently, the time of the local exposure to the epidemics is one of the critical parameters of the impact, so the case data was adjusted and aligned at a similar phase in the development of the epidemics, chosen based on the availability of data at approximately, local Time Zero + two months, i.e. approximately two months after the first local exposure to the infection. In the study this translates to the beginning of April, 2020 for Wave 1 cases (LTZ in January, 2020) and beginning of May for Wave 2 (LTZ end of February to early March, 2020).

A combined dataset of approximately forty cases was thus constructed based on the conditions outlined in [2], essentially, bringing together the cases with similar social and economic parameters to minimize the number of potentially influencing factors along with the expectation of certain minimal level of exposure to the epidemics and reliability of the reported data.

The dataset was constructed from the publicly available current data on the epidemics impact per case, i.e., reporting jurisdiction. It comprises the current value of the epidemics impact recorded in the jurisdiction (case) and measured in in mortality per capita *m(t)* (M.p.c.), per million of population, and a number of observable parameters selected as described further in this section with the hypothesis of a certain level of correlation between the observable parameter set and the severity of the outcome.

On the relative scale of impact by jurisdiction, the “explosive” cases were normally identified as those with relative M.p.c. (relative to the maximum among all reporting jurisdictions worldwide) of around and above 0.5. This subgroup of cases included all commonly reported cases of high epidemics impact at the time of writing.

In evaluation of distribution in the coordinates of principal components two higher impact clusters of cases were identified by relative impact: explosive cases with relative M.p.c. above 0.8 group included the well-known first wave cases: Italy; Spain and New York with the highest impact worldwide observed to date. In the second group were six somewhat milder-impact cases, namely: United Kingdom; France; Belgium; Netherlands; Ireland and Quebec (Canada), with relative M.p.c. in the range from 0.6 to 0.8.

The impact parameter was not used in the training of the unsupervised learning models (i.e. excluded from the training dataset) but only for identification of the regions of interest (i.e. higher epidemiological impact) in the latent representations produced by the models as a result of training.

### 2.2 Observable Parameters

The examples of factors of influence can include, among others: genetic differences; population density, social traditions and cultural practices, past widespread public policy such as immunization; smoking habits and of course the epidemiological policy of the jurisdiction aimed at controlling the spread of the disease.

In addition to the common measurable factors such as population density, age demographics, smoking prevalence a number of additional factors with potential impact on the severity of the epidemics pattern were considered in this study as described in this section. A common comment for some of them is that due to limitation of time and resources, a rating scale approach was chosen for those factors that cannot or would be challenging to measure directly. Understandably, such an approach can be influenced by subjective perceptions; however, we believe that more robust and objective techniques can be developed over time improving the quality of the analysis and the resulting conclusions.

#### Connectivity

intended to measure the intensity of international and regional connections in the jurisdiction of the case, for example, international, inter and intra-regional travel and migration, tourism; seasonal and work-related migration and so on; more intensive connection hubs can be expected to have higher exposure to the pandemics increasing the probability of a heavier impact.

#### Social proximity

intended to reflect the closeness of inter-personal connections in the case, again in multiple spheres and domains, for example: family connections; socializing practices and traditions; the intensity of business connections; lifestyle practices; social events and others. Again, as was commented previously modeling such a complex factor as a single value parameter may open the analysis to the vulnerability of subjectiveness; yet we believed that it could be important for the analysis and improvements to make its evaluation, by case more objective and accurate are possible in the future studies.

We also used three rating parameters intended to measure the policy of the jurisdiction as relates to the response to the pandemics. They are: 1) epidemiological preparedness of the public healthcare system to an intensive and rapid development of an epidemics; 2) the effectiveness of the policy response; and 3) the timeliness of the response.

#### Epidemiological preparedness

intended to measure the preparedness of the health care system to handle a rapid onset of a large-scale epidemics. This parameter is intended to be specific to epidemiological situation rather than the general state of the health care system, its technological level, funding and so on).

#### Policy response

intended to indicate the quality of the public health policy in controlling the epidemics based on available scientific data at the time including its clarity and availability for understanding and following by the general population facilitating its preparedness to participate. While some concerns can be expressed that this factor can be influenced by post-impact considerations with potential post-factum correlated with the outcome, we believe that with the accurate approach these risks can be minimized. For example, it is evident that an unclear or misleading policy message could be highly detrimental to the intended effect and one doesn’t need the outcome to judge such policy parameters objectively at the time the decision is made and before the outcome is recorded.

#### Policy timeliness

measures the relative timing of introduction of the epidemiological policy to the local exposure and development of the epidemics.

#### Universal BCG immunization record

indicates the record of a current or previous immunization program according to classification introduced in [9]. A detailed definition is provided in the Appendix.

#### Epidemiological impact

measured in Covid-19 caused mortality per 1 million capita relative to the world’s maximum value at the time of the analysis.

The resulting dataset of 40 national and subnational cases with the identified observable parameters and the recorded epidemiological impact at the time of preparation is presented in Table 1, Appendix.

**Table 1.** Linear Regression Analysis.

| Factor | Linear Regression | Correlation |
| --- | --- | --- |
| Policy, Timing | 0.534 | 0.906 |
| Connection hub | 0.196 | 0.697 |
| Policy, Effectiveness | 0.094 | 0.856 |
| BCG immunization | 0.092 | 0.686 |
| Social proximity | 0.078 | 0.794 |

### 2.3 Machine Learning Methods

To evaluate the hypothesis of the correlation between the identified parameters and the epidemiological outcome in the case, several well-known machine learning methods were used:

1. Linear regression.
2. Principal Component Analysis and identification of principal informative factors.
3. Unsupervised deep neural network-based dimensionality reduction and selection of dominant informative factors.

The first method produces a best fit linear approximation of the resulting effect series with a total deviation (error) from the trend [10].

Principal Component Analysis [7] produces a linear transformation of the dataset to the coordinates with the highest variation and does not use the resulting effect labels.

A deep neural network autoencoder (method 3) produces a non-linear dimensionality reduction of the observable data to the lower-dimensional representation with the most informative features [8]. The structure of the deep neural network model used in this work is described in detail in [11]. The diagram of the architecture of the unsupervised autoencoder model is given in Fig.1.

**Fig. 1.**
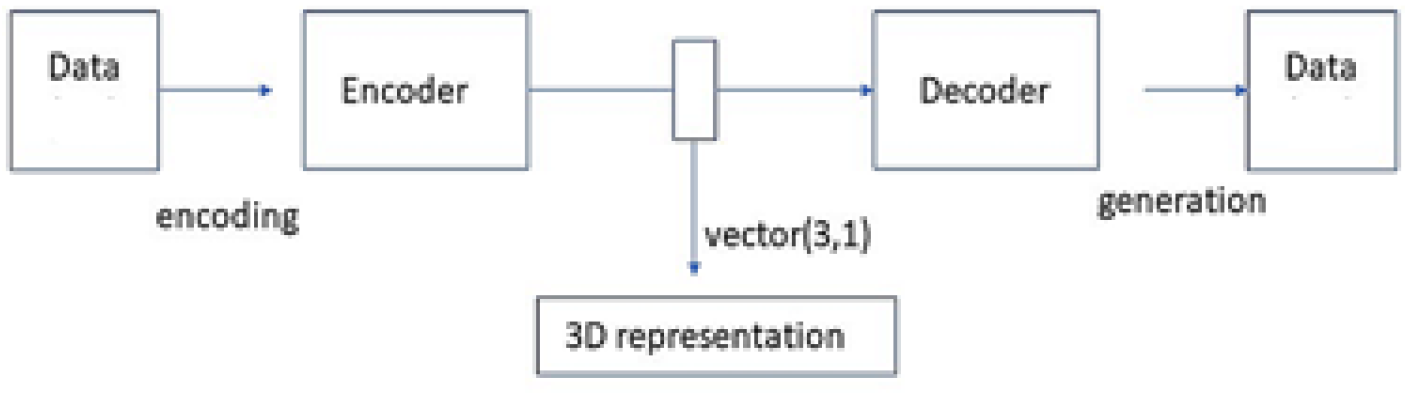
Dimensionality reduction with deep autoencoder.

In the unsupervised training phase, the model is trained to reproduce the input data with good accuracy and thus does not require labels marked with the outcome; the same applies to Method 2 (PCA). Achieving an improvement in the accuracy of reproduction of the input data, that can be measured by a number of training metrics indicates that the model has learned some essential characteristics of the initial distribution. Once trained, the unsupervised model can perform two essential transformations: the encoding one, from the observable data creating a representation image; and the generative one, from the representation to the observable data space. The aim of unsupervised learning is thus to minimize the deviation of the original training sample from its regeneration created by the model.

The models were implemented in Python, Keras, Tensorflow [12] with a number of common machine learning and data processing packages.

## 3 Results

### 3.1 Linear Regression

Linear regression with 8 identified input parameters produces a trend with a strong match score to the label of *0*.*9* of out 1.0 maximum. The most influential factors in the regression trend are shown in Table 1.

Policy factors were expected to have a strong influence on the outcome of the case that is confirmed by the results of the linear regression analysis. As well, the importance of other factors such as connection intensity, social proximity culture, BCG immunization and smoking was observed.

### 3.2 Principal Component Analysis

Principal component analysis identified three principal components with overall influence of 96% as described in Table 2. The highest influence factors in the PCA analysis are mostly aligned with the results of the linear regression analysis: policy-time, connection hub, social proximity, BCG and the smoking rate.

**Table 2.** Principal Components.

| Eigenvector | Main factors | Relative Weight |
| --- | --- | --- |
| Axis 1 | Policy-time, BCG | 0.570 |
| Axis 2 | BCG, smoking | 0.166 |
| Axis 3 | Connection hub, social proximity | 0.127 |

PCA transformation is inherently unsupervised method of learning, meaning that the prior known outcome labels are not required to learn the principal components as well as representation of the input data in the coordinates of identified principal component eigenvectors. By plotting thus transformed dataset in the coordinates of the principal component vectors, interesting result can be observed by identifying the cases with the highest impact of the epidemics.

Shown in Fig.1 are the visualizations of the distribution of the dataset in the coordinates of principal components identified by PCA analysis. The cases and the resulting region of the highest-impact cluster is shown in blue; whereas the milder cluster of 6 cases, in magenta.

In the visualizations of the case clusters in the principal component representation one can observe a clear separation of the higher-impact case clusters from the general background cases. Such a clear separation allows to identify the region where the cases with potentially higher impact including the “explosive” pattern can be located, in the coordinates of principal component representation as well as in the initial, observable parameter space, with the possibility to identify the combinations of the observable parameters that can be linked to higher impact outcomes.

### 3.3 Unsupervised Autoencoder Model

A similar approach to the previous section can be demonstrated with an unsupervised neural network autoencoder model that reduces the number of parameters by compressing the observable data space into a lower dimensional representation while unsupervised training process is aimed at improving the accuracy of regeneration form the compressed representation to the observable space. Models of a similar type were used to create structured unsupervised representations of different data types via unsupervised autoencoder training with minimization of generative error [8,11].

The dimensionality of the unsupervised representation for the models in the study that is defined by the size of its central encoding layer was chosen based on the results of the Principal Component Analysis in the previous section, indicating three most informative components.

Presented in Fig.3 are direct visualizations of the distributions of data in the unsupervised representation created by a trained autoencoder model.

**Fig. 2.**
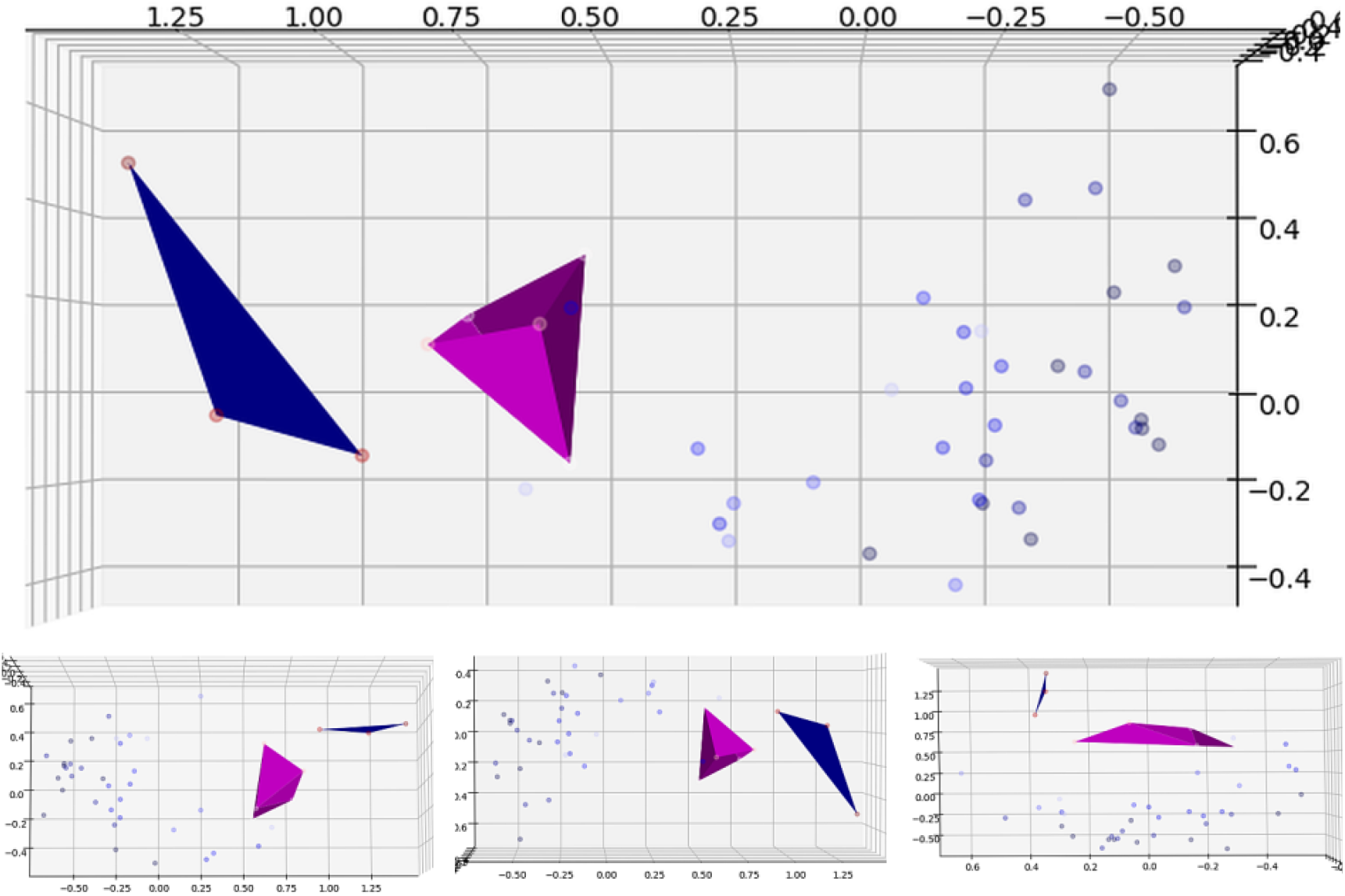
High-impact cluster identification with PCA.

**Fig. 3.**
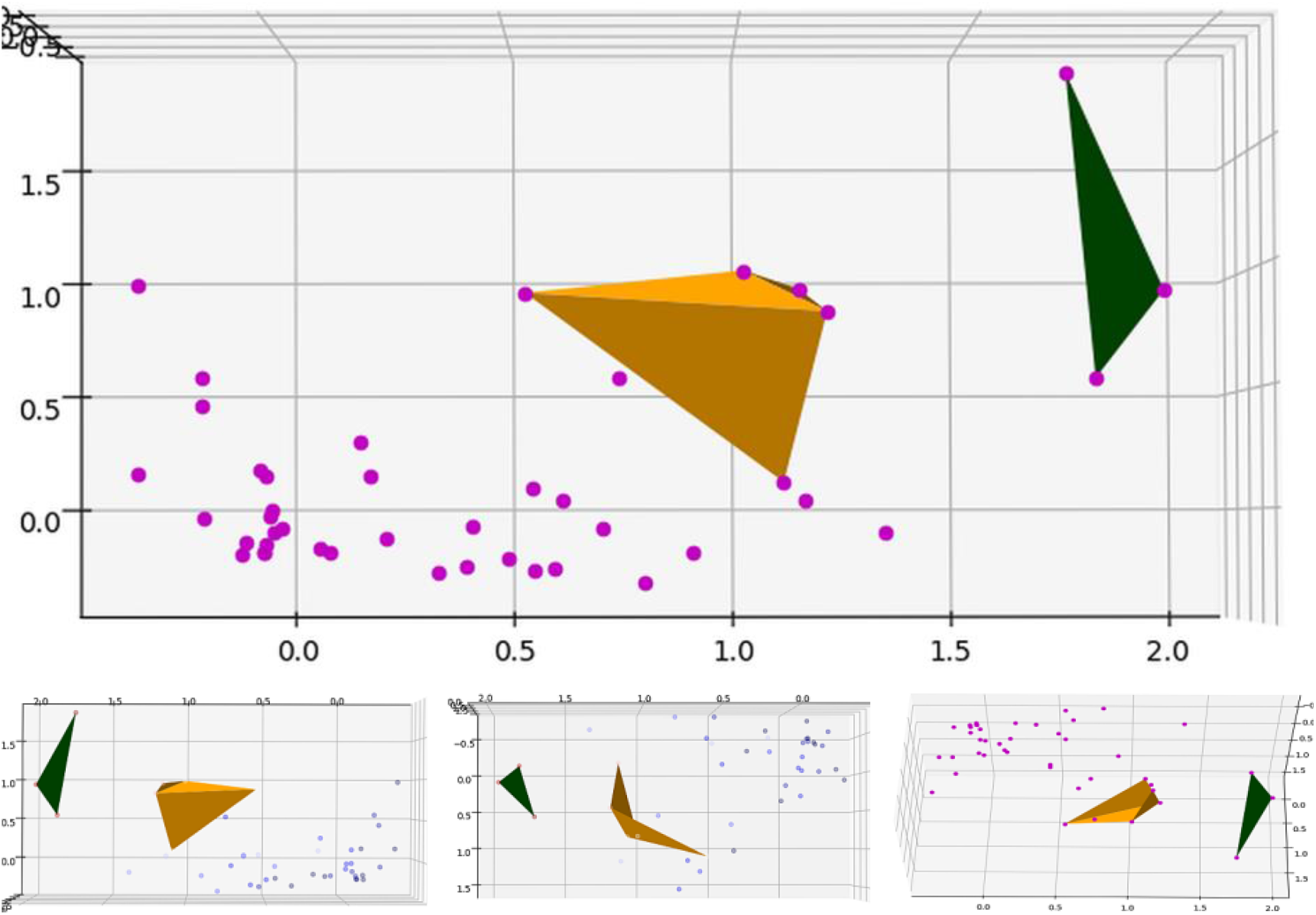
High-impact clusters in the autoencoder representation.

The highest impact cluster (3 cases) is shown in Fig.3 in green whereas the milder one (6 cases), in orange. Again, a similar pattern of clear separation of higher-impact cases from general background can be observed with these models, in agreement with the results of PCA analysis in the previous section.

It is worth noting that as with PCA in the previous section, autoencoder models though essentially non-linear, also allow to identify the higher-impact regions in the coordinates of the observable parameters. This can be achieved by forward-propagating through the generative part of the model the identified region of interest, defined by a set of characteristic points in the latent representation, thus allowing to identify the image of the region of interest in the latent representation coordinates in the observable space. The combinations of observable parameters that produce the effect of interest or concern can thus be identified proactively, and used in development of an effective epidemiological policy.

## 4 Conclusion

The methods of unsupervised machine learning can be effective in identifying and separating the informative features in complex general data [13,14]. In this work, two different methods of unsupervised learning applied independently, consistently demonstrated good separation of cases with higher Covid-19 epidemiological impact from the general background.

The analysis and the findings of the study can be used in evaluation of possible epidemiological scenarios in jurisdiction based on evaluation of the factors identified and discussed in this work, as well as those that can be added in the subsequent studies. Further research and development in the identified direction has a potential of producing effective modeling tools to identify the areas of potential epidemiological risk in the public healthcare policy and design corrective and / or preventative measures to avoid the heavier impact scenarios.

Further studies can be focused on improving the accuracy of measurement of the identified observable parameters as well as introducing additional ones, leading to higher accuracy and confidence of the evaluation.

## Data Availability

The data referred to is enclosed or referenced in the manuscript

## Appendix Case Dataset

**Table 1.** Combined Case Dataset, at LTZ + 2m. Sources: [15 – 18] and others

| *Case* | *Policy* |  |  | *Conn* | *Bcg* | *Smo* | *Den* | *Soc* | *Age* | *Impact* |
| --- | --- | --- | --- | --- | --- | --- | --- | --- | --- | --- |
|  | *p-prep* | *p-qlty* | *p-tme* |  |  |  |  |  |  |  |
| Taiwan | 0 | 0 | 0 | 0.1 | 0 | 0.34 | 0.3 | 0.2 | 0.3 | 0.001 |
| Japan | 0.1 | 0.1 | 0 | 0.6 | 0 | 0.674 | 0.3 | 0.2 | 0.5 | 0.002 |
| Singapore | 0 | 0 | 0 | 0.4 | 0 | 0.33 | 0.5 | 0.3 | 0.25 | 0.004 |
| Australia | 0.2 | 0.2 | 0 | 0.2 | 0.3 | 0.298 | -0.5 | 0.3 | -0.4 | 0.005 |
| South Korea | 0.1 | 0.2 | 0 | 0.2 | 0 | 0.996 | 0.3 | 0.2 | 0 | 0.013 |
| Finland | 0.3 | 0.2 | 0.1 | 0.1 | 0.3 | 0.418 | -0.2 | 0.2 | 0.3 | 0.017 |
| Canada | 0.4 | 0.2 | 0.2 | 0.3 | 0.8 | 0.354 | -0.5 | 0.4 | 0 | 0.023 |
| Ontario (Canada) | 0.4 | 0.2 | 0.25 | 0.3 | 0.8 | 0.258 | -0.2 | 0.4 | 0 | 0.025 |
| Germany | 0.3 | 0.2 | 0.2 | 0.5 | 0.2 | 0.608 | 0.2 | 0.4 | 0.5 | 0.052 |
| Sweden | 0.3 | 0.3 | 0.3 | 0.1 | 0.6 | 0.412 | 0.0 | 0.3 | 0 | 0.148 |
| UK | 0.5 | 0.7 | 0.7 | 0.7 | 0.8 | 0.398 | 0.2 | 0.5 | 0 | 0.248 |
| France<sup>1</sup> | 0.5 | 0.5 | 0.6 | 0.7 | 0.6 | 0.596 | 0.2 | 0.7 | -0.2 | 0.371 |
| Belgium | 0.5 | 0.4 | 0.5 | 0.7 | 1 | 0.53 | 0.2 | 0.5 | 0 | 0.429 |
| Spain | 0.8 | 0.7 | 0.8 | 0.5 | 0.8 | 0.584 | 0.2 | 0.8 | 0.5 | 0.965 |
| Italy | 0.8 | 0.8 | 0.9 | 0.7 | 1 | 0.566 | 0.2 | 0.8 | 0.5 | 0.969 |
| USA | 0.5 | 0.5 | 0.5 | 0.3 | 1 | 0.39 | -0.2 | 0.4 | -0.4 | 0.095 |
| New York (USA) | 0.8 | 0.8 | 0.9 | 1 | 1 | 0.25 | 0.5 | 0.8 | -0.5 | 1.000 |
| California (USA) | 0.5 | 0.3 | 0.2 | 0.5 | 1 | 0.226 | 0.1 | 0.4 | -0.5 | 0.040 |
| Slovakia | 0.2 | 0.2 | 0.2 | 0 | 0 | 0.794 | 0.2 | 0.2 | -0.1 | 0.016 |
| Argentina | 0.4 | 0.3 | 0.3 | 0 | 0 | 0.478 | -0.2 | 0.3 | -0.5 | 0.019 |
| Chile | 0.2 | 0.2 | 0.1 | 0 | 0 | 0.76 | 0.1 | 0.2 | -0.5 | 0.050 |
| Ukraine | 0.6 | 0.4 | 0.3 | 0 | 0 | 0.94 | 0.2 | 0.4 | 0.1 | 0.027 |
| Poland | 0.3 | 0.2 | 0.1 | 0.2 | 0 | 0.648 | 0.2 | 0.3 | -0.1 | 0.066 |
| Moldova | 0.6 | 0.4 | 0.3 | 0 | 0 | 0.56 | 0.2 | 0.4 | -0.4 | 0.125 |
| Czechia | 0.3 | 0.2 | 0.1 | 0.1 | 0 | 0.766 | 0.2 | 0.25 | 0 | 0.082 |
| Croatia | 0.3 | 0.2 | 0.1 | 0 | 0 | 0.74 | 0.2 | 0.25 | 0.5 | 0.068 |
| Albania | 0.3 | 0.2 | 0.1 | 0 | 0 | 0.8 | 0.2 | 0.25 | -0.5 | 0.038 |
| Greece | 0.2 | 0.1 | 0 | 0.4 | 0 | 1 | 0.2 | 0.5 | 0.5 | 0.049 |
| Israel | 0.1 | 0.1 | 0.1 | 0.4 | 0.3 | 0.382 | 0.2 | 0.2 | -0.5 | 0.094 |
| Prairies<sup>(1)</sup> (Canada) | 0.3 | 0.2 | 0.2 | 0 | 0.6 | 0.292 | -0.3 | 0.2 | -0.3 | 0.016 |
| Quebec (Canada) | 0.6 | 0.4 | 0.5 | 0.3 | 0.8 | 0.304 | -0.2 | 0.5 | 0.3 | 0.912 |
| Norway | 0.2 | 0.2 | 0.2 | 0.2 | 0.2 | 0.452 | -0.2 | 0.25 | -0.1 | 0.138 |
| Denmark | 0.2 | 0.2 | 0.2 | 0.1 | 0.3 | 0.352 | 0.2 | 0.25 | 0.1 | 0.303 |
| Switzerland | 0.2 | 0.2 | 0.2 | 0.2 | 0.3 | 0.51 | 0.2 | 0.25 | 0.25 | 0.603 |
| Austria | 0.2 | 0.2 | 0.2 | 0.2 | 0.2 | 0.704 | 0.2 | 0.25 | 0.4 | 0.238 |
| Portugal | 0.3 | 0.3 | 0.3 | 0.3 | 0 | 0.63 | 0.2 | 0.5 | 0.5 | 0.355 |
| Ireland<sup>(2)</sup> | 0.4 | 0.3 | 0.5 | 0.4 | 0.2 | 0.444 | 0.2 | 0.6 | -0.4 | 0.653 |
| Netherlands | 0.3 | 0.4 | 0.4 | 0.5 | 1 | 0.524 | 0.3 | 0.25 | 0.4 | 0.774 |

<sup>1</sup> Manitoba and Saskatchewan provinces, Canada
<sup>2</sup> Inconsistencies in implementation of universal BCG policy, [19]

### Observable factors

#### Policy

p-prep: health care preparedness, range 0 .. 1, lower to higher preparedness

p-qlty: response measures, range 0 .. 1, lower to higher epidemiological policy quality;

p-tme: response timing, range 0 .. 1, timely to delayed

**Conn:** connection intensity, range 0 .. 1, lower to higher connection intensity

**Bcg:** BCG immunization record, range 0 .. 1. The value of 0 indicates current or very recent universal immunization policy; the value of 1 indicates no effective immunization policy and equivalent cases [2]. A value between 0 and 1 indicates a previous universal immunization policy relative to the time after cessation.

**Smo:** smoking prevalence in the population. In the cases with large disparity between genders and so on, the higher of values was taken.

**Den:** population density. Due to significant variability in population density between the cases in the dataset, a logarithmic band scale was used; additionally, in cases with very large territory, a negative offset was added to account for non-homogeneousness of the distribution of individual cases and the delay in propagation of the epidemics due to geographical distance, as illustrated in the examples in Table 2 (below). A higher granularity analysis of national jurisdictions with very high geographical spread can be attempted in a future study.

**Age:** age demographics, median age, logarithmic band of the deviation from the dataset mean, range: −0.5 .. 0.5.

### Outcome factors

Impact: the epidemiological impact in the jurisdiction at the time of analysis measured as relative mortality per 1 million capita (R.mpc), relative to world’s highest at the time.

### Population density

#### Table Population density evaluation

| *Case* | *Overall density* | *Logarithmic band* | *Distance offset* | *Factor value* |
| --- | --- | --- | --- | --- |
| New York City (USA) | 10431 | 0.5 | 0 | 0.5 |
| Taiwan | 671 | 0.3 | 0 | 0.2 |
| Japan | 348 | 0.3 | 0 | 0.3 |
| Singapore | 8358 | 0.5 | 0 | 0.5 |
| Australia | 3.2 | 0 | - 0.5 | - 0.5 |
| Finland | 18.2 | 0.1 | - 0.3 | - 0.2 |
| Canada | 4 | 0 | - 0.5 | - 0.5 |
| Ontario (Can) | 18.2 | 0.1 | - 0.3 | - 0.2 |
| Prairies (Can) | 2 | 0 | - 0.3 | - 0.3 |
| USA | 36.0 | 0.2 | -0.4 | - 0.2 |

## References

1. Miller A., Reandelar M-J., Fasciglione K., Roumenova V., Li Y., Otazu G.H. Correlation between universal BCG vaccination policy and reduced morbidity and mortality for COVID-19: an epidemiological study, medRxiv 2020.03.24.20042937 (2020).

2. Dolgikh S., Further evidence of a possible correlation between the severity of Covid-19 and BCG immunization, preprint MedRxiv, https://www.medrxiv.org/con-tent/10.1101/2020.04.07.20056994v1 April 2020.

3. Sharma A., Sharma S.K., Shi Y., et al. BCG vaccination policy and preventive chloroquine usage: do they have an impact on COVID-19 pandemic? Cell death & disease, 11(7), 1–10 (2020).

4. Yitbarek K., Abraham G., Girma T. et al. The effect of Bacillus Calmette–Guérin (BCG) vaccination in preventing sever infectious respiratory diseases other than TB: implications for the COVID-19 pandemic. Vaccine 38(41), 2020, 6374–6380 (2020).

5. Ebina-Shibuya, R., Horita, N., Namkoong, H., Kaneko, T. National policies for paediatric universal BCG vaccination were associated with decreased mortality due to COVID-19. Respirology (Carlton, Vic.), http://europepmc.org/article/pmc/pmc7323121 (2020).

6. Dayal, D., Gupta, S. Connecting BCG vaccination and COVID-19: additional data. Medrxiv https://www.medrxiv.org/content/medrxiv/early/2020/04/19/2020.04.07.20053272.full.pdf (2020).

7. Jolliffe I.T., Principal Component Analysis, Series: Springer Series in Statistics, 2nd ed, Springer, NY 2002.

8. Bengio Y., Learning deep architectures for AI, Foundations and Trends in Machine Learning, vol.2, no.1, 1–127 (2009).

9. Zwerling A., Behr M.A., Verma A., Brewer T.F., Menzies D., Pai M. The BCG World Atlas: A Database of Global BCG Vaccination Policies and Practices PLOS Medicine https://doi.org/10.1371/journal.pmed.1001012 (2011).

10. Freedman D., Statistical Models: Theory and Practice. Cambridge University Press, 2005.

11. Prystavka P., Cholyshkina O., Dolgikh S., Karpenko D. Automated object recognition system based on aerial photography. In: 10th International Conference on Advanced Computer Information Technologies ACIT-2020 Deggendorf, Germany (in print) (2020).

12. Keras: the Python deep learning API https://keras.io/

13. Polat K., Gunes S. An expert system approach based on principal component analysis and adaptive neuro-fuzzy inference system to diagnosis of diabetes disease, Digital signal processing, 17, 702–710, (2007).

14. Zhang Z., Costell A. Principal components analysis in clinical studies. Annals of Translational Medicine, 5 (17), 351 (2017).

15. BCG World Atlas http://www.bcgatlas.org/ (4.04.2020).

16. Coronavirus map Google https://www.google.com/covid19-map/ (4.04.2020).

17. World smoking prevalence https://ourworldindata.org/smoking (4.04.2020).

18. Word population data https://www.worldometers.info/world-population (4.04.2020).

19. Sweeney E., Dahly D., Seddiq N., et al. Impact of BCG vaccination on incidence of tuberculosis disease in southern Ireland, BMC Infectious Diseases, 19. 397 (2019).

